# Sex-specific Trends in Incidence, Prevalence, and Mortality of Chronic Kidney Disease (CKD) in the United States, 1990-2019: Findings from the Global Burden of Disease Study

**DOI:** 10.1101/2025.10.06.25337332

**Authors:** Nam Nguyen

## Abstract

**Background:** Chronic kidney disease (CKD) remains a major public health burden in the United States, affecting over 35 million adults. Despite improvements in the management of hypertension and diabetes, sex-based disparities in CKD burden persist, with women exhibiting higher prevalence but men facing worse outcomes.

**Objective:** This study examined sex-specific trends in the incidence, prevalence, and mortality of CKD in the United States from 1990 to 2019 using data from the Global Burden of Disease (GBD) 2021 database.

**Methods:** A retrospective analysis was conducted using age-standardized CKD incidence, prevalence, and mortality rates per 100,000 population, stratified by sex. Independent samples t-tests and Levene’s tests evaluated sex differences, while effect sizes (Cohen’s d, Hedges’ g, and Glass’s Δ) quantified their magnitude. Linear regression models assessed temporal trends over the 29-year period.

**Results:** Females exhibited significantly higher mean incidence (326.10 vs. 290.34 per 100,000; P < 0.001) and prevalence (8,115.47 vs. 6,726.53 per 100,000; P < 0.001) than males, with large effect sizes across all measures from 1990 to 2019. Conversely, males demonstrated higher mean mortality (16.94 vs. 12.35 per 100,000; P < 0.001). Temporal analyses revealed modest increases in incidence for both sexes, a significantly faster rise in prevalence among females (β = 22.46, P < 0.001), and higher mortality growth among males (β = 0.57, P < 0.001).

**Conclusions:** From 1990 to 2019, CKD incidence and prevalence increased more rapidly among women, whereas men consistently experienced higher mortality. These findings highlight persistent and widening sex disparities in CKD burden, likely reflecting both biological differences and gender-related inequities in healthcare access. Future prevention and treatment strategies should incorporate sex-specific approaches to reduce progression and mortality from CKD.

## Introduction

Every year, chronic kidney disease (CKD) poses a strong health burden in the United States, affecting more than 1 in 7 adults, an estimated 35 million people, and ranks consistently among the top ten causes of death [1,2]. CKD arose mainly from hypertension, diabetes, and aging, where about 1 in 3 adults with diabetes and 1 in 5 adults with high blood pressure were also affected [3,4]. As the United States population continues to grow older, the prevalence of CKD is also projected to rise further, since more than one-third of the adults aged 65 years and older already live with the disease compared to only about 6% of the adults aged 18-44 [1]. Even in its early stages, CKD can influence the risk of cardiovascular disease, hospitalization, and premature mortality, placing a strain on patients and the health system [5].

Sex differences have shown to complicate this burden. Even though it has shown that CKD prevalence is higher for women, men tend to experience faster progression of this disease, are about 60% more likely to develop end-stage kidney disease (ESKD) and face higher mortality [1,6,7]. From the literature, biological factors (e.g., hormonal differences and renal hemodynamics) and gender-related factors (e.g., health behaviors and patterns of treatment initiation) are believed to contribute to these disparities [7,8]. Despite current advances in the management of hypertension and diabetes, CKD incidence and mortality have remained highly persistent across the past three decades, emphasizing a need for more nuanced surveillance and intervention strategies [1,2].

By understanding the sex-specific differences in chronic kidney disease burden, hospitals can tailor more nuanced prevention and treatment efforts, reducing the progression to kidney failure and close the gaps in outcomes. This study examines the temporal trends in incidence, prevalence, and mortality rates of CKD in the United States from 1990 to 2019, with a focus on sex differences utilizing age-standardized data from the Global Burden of Disease (GBD) database.

## Methods And Materials

This was a retrospective study analyzing the changes in incidence, prevalence, and mortality of CKD, in terms of sex, in the United States from 1990 to 2019, using data from the GBD 2021 database. The GBD database provided age and sex specific disease burden estimates. The data were aggregated by, and age-adjusted rates of incidence, prevalence, and mortality were all calculated for males and females separately, with presented as rates per 100,000 population. These rates were selected to adequately assess the dynamics of sex-differentiated CKD results.

The study population was defined as the residents of the United States, where the primary outcome variables were standardized rates of chronic kidney disease. The GBD carefully explores the methods for calculating injury rates, utilizing a myriad of data resources such as hospital admissions and discharges, cause-of-death certificates, and household forms completed by the relatives of the deceased. Then, this data is further processed into the national dataset. For this specific analysis, the age-standardized CKD rates for both males and females were aggregated every year between 1990 and 2019. Additionally, exploratory statistical methods were utilized to describe the distribution of CKD and inferential statistics to analyze CKD variability dependent on sex. An independent sample t-test test was also employed to determine any sex differences in the mean CKD incidence, prevalence, and mortality rates. Levene’s test was applied before each *t*-test to assess whether the assumption of equal variances had been met. Measures of the effect sizes (i.e., Cohen’s Δ, Hedges’ g, and Glass’s D) were analyzed to quantify the magnitude of the observed sex differences. In short, these effect size measurements were included to understand the sex differences on a practical context than just relying on the statistical significance of the *t*-tests.

Temporal trend analysis was conducted to evaluate the change in CKD rates over the study period. Linear regression models were also used to measure incidence, prevalence, and mortality rates, utilizing as the independent. Separate models were also measured for males and females to identify differences in the rates of CKD decline or stabilization. Moreover, 95% confidence intervals were calculated to measure the precision of the estimates, and all statistical tests were conducted with a P value threshold of <0.05. Additionally, the analysis was performed utilizing statistical software, with the representations focusing on the temporal dynamics and sex differences in CKD outcomes over the 29-year period.

## Results

The study analyzed the sex-specific trends in incidence, prevalence, and mortality rates of CKD in the United States from 1990 to 2019 utilizing the GBD database. The data was stratified by sex, where age-standardized rates per 100,000 population were calculated for each year. Statistical analyses revealed significant differences in CKD outcomes between males and females, where females demonstrate higher rates of incidence and prevalence; meanwhile, males had higher rates of mortality.

### Incidence

The incidence of CKD showed significant sex differences [Figure 1]. Females had a higher mean incidence rate (326.10 per 100,000, standard deviation [SD] = 7.21) compared to males (290.34 per 100,000, standard deviation [SD] = 6.76). The independent samples *t*-test indicated a significant difference in the incidence rates between the sexes (*t*[58] = −19.820, *P* < 0.001). Effect size of the metrics also details the magnitude of this difference, with Cohen’s d calculated at 6.989, Hedge’s g at 6.989, and Glass’s Δ at 7.212 [Table 1]. These large effect sizes illustrate that the disparity in CKD incidence rate between males and females was substantial.

**Table 1.**
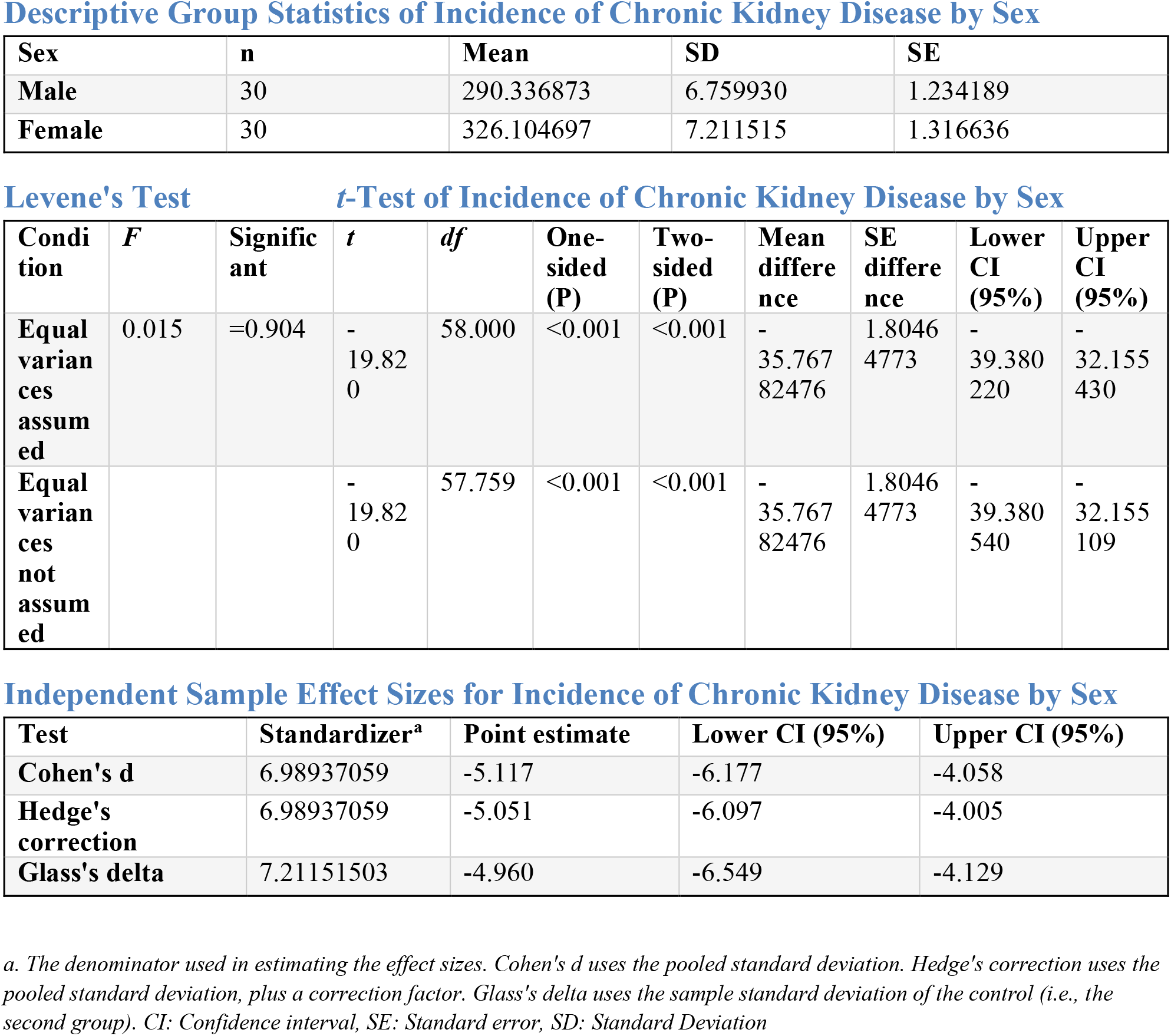

**Figure 1:**
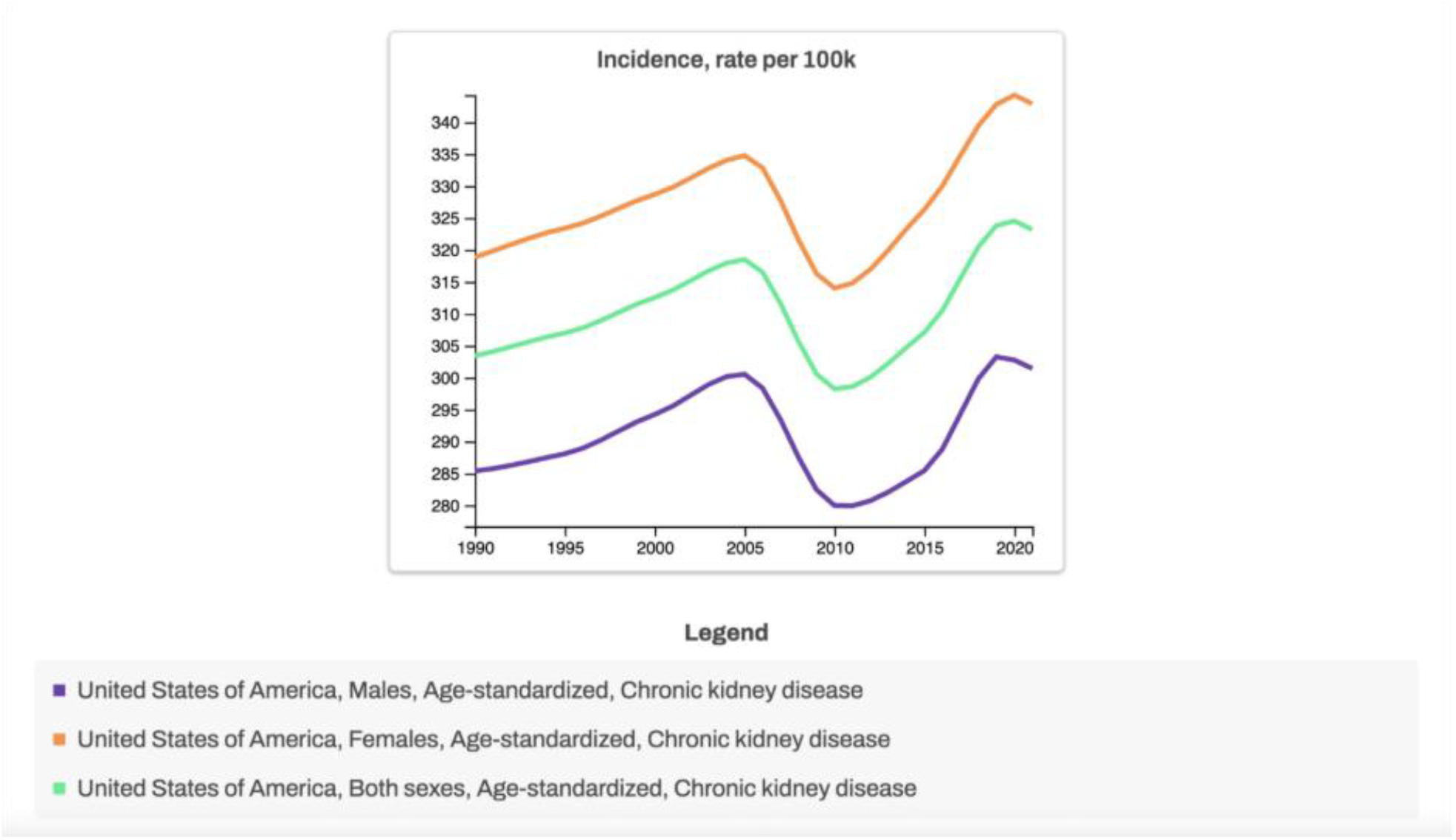

Over the 27-year period, the incidence of CKD for both sexes increased, yet the pattern of increase differed slightly. In 1990, the incidence rate for males was 285.37 per 100,000, increasing to 303.25 per 100,000 by 2019, a 6.1% increase. For females, the incidence rate in 1990 was 318.87 per 100,000, which slightly increase to 342.78, representing 8.1% increase. Although both sexes experienced increases in incidence, females consistently exhibited higher amounts of incidence rates than males throughout the study period.

The temporal trend annual incidence illustrated no significant linear trend in either sex (males: β = 0.063 per year, 95% CI −0.234 to 0.359, p = 0.668; females: β = 0.256 per year, 95% CI −0.046 to 0.557, p = 0.093) [Table 1].

### Prevalence

The prevalence of CKD showed significant sex differences [Figure 2]. Females had a higher mean prevalence rate (8115.47 per 100,000, standard deviation [SD] = 242.62) compared to males (6726.53 per 100,000, standard deviation [SD] = 130.34). The independent samples *t*-test indicated a significant difference in the prevalence rates between the sexes (*t*[58] = −27.622, *P* < 0.001). Effect size of the metrics also details the magnitude of this difference, with Cohen’s d calculated at 194.75, Hedge’s g at 194.75, and Glass’s Δ at 242.62 [Table 2]. These large effect sizes illustrate that the disparity in CKD prevalence rate between males and females was substantial.

**Table 2.**
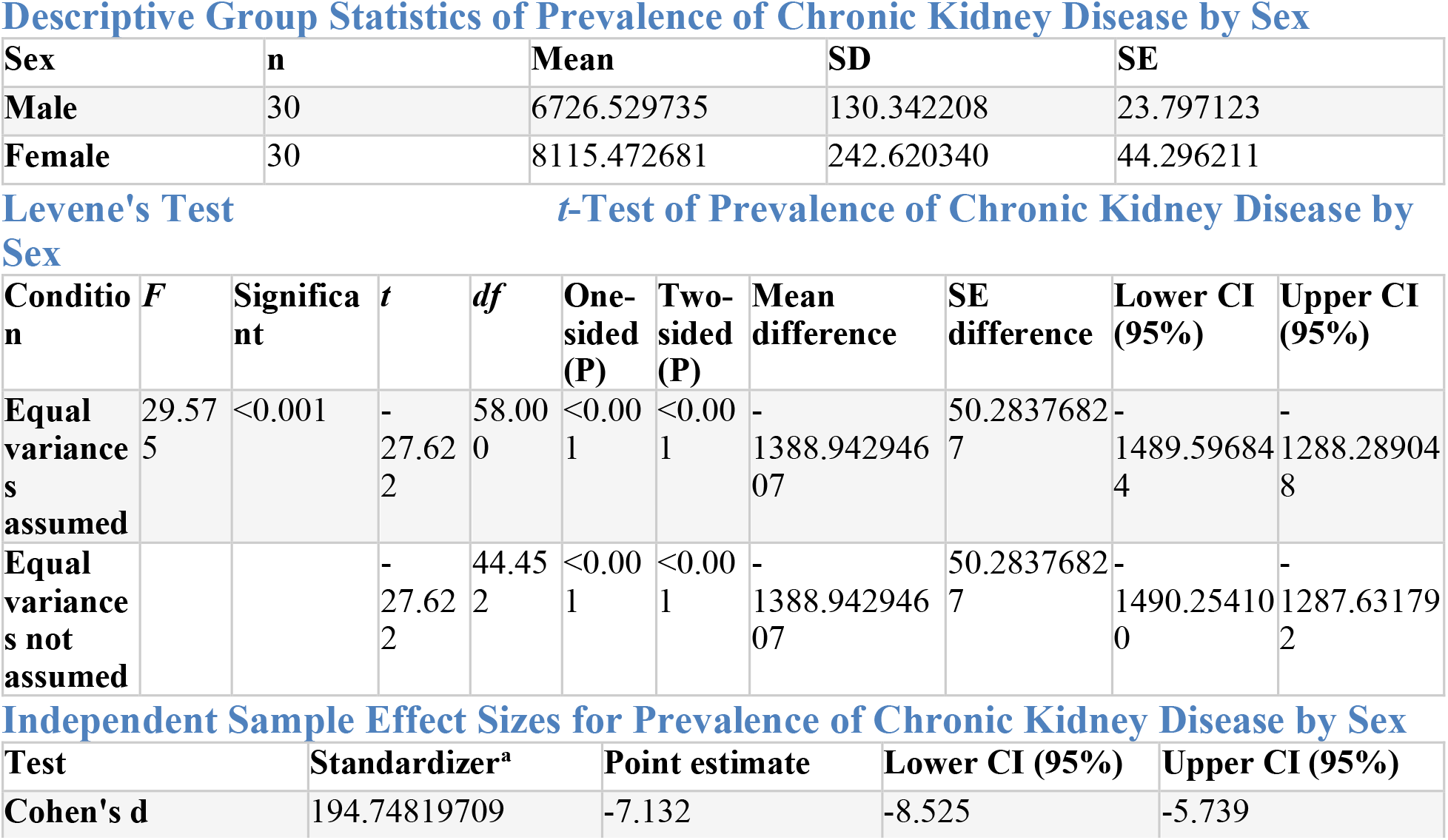
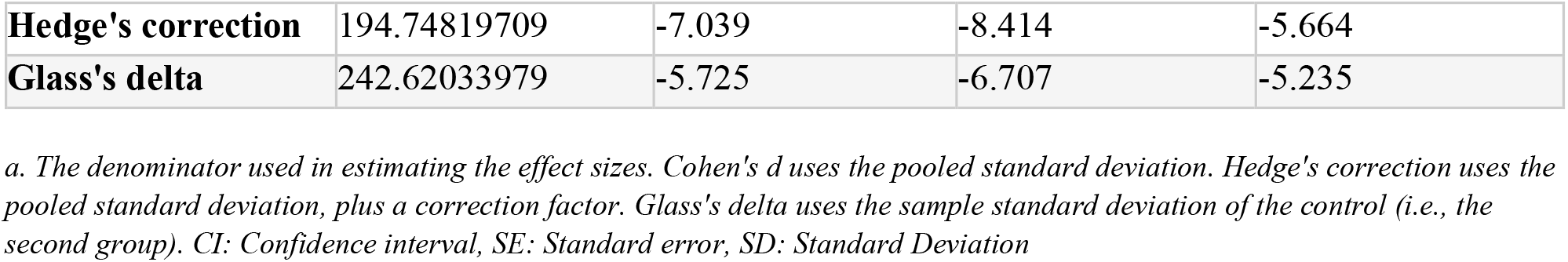
Descriptive Group Statistics of Prevalence of Chronic Kidney Disease by Sex

| Sex | n | Mean | SD | SE |
| --- | --- | --- | --- | --- |
| Male | 30 | 6726.529735 | 130.342208 | 23.797123 |
| Female | 30 | 8115.472681 | 242.620340 | 44.296211 |

**Levene's Test** ***t*-Test of Prevalence of Chronic Kidney Disease by Sex**
| Condition | *F* | Significant | *t* | *df* | One-sided (P) | Two-sided (P) | Mean difference | SE difference | Lower CI (95%) | Upper CI (95%) |
| --- | --- | --- | --- | --- | --- | --- | --- | --- | --- | --- |
| Equal variances assumed | 29.575 | <0.001 | -27.622 | 58.000 | <0.001 | <0.001 | -1388.94294607 | 50.28376827 | -1489.596844 | -1288.289048 |
| Equal variances not assumed |  |  | -27.622 | 44.452 | <0.001 | <0.001 | -1388.94294607 | 50.28376827 | -1490.254100 | -1287.631792 |

**Independent Sample Effect Sizes for Prevalence of Chronic Kidney Disease by Sex**
| Test | Standardizer<sup>a</sup> | Point estimate | Lower CI (95%) | Upper CI (95%) |
| --- | --- | --- | --- | --- |
| Cohen's *d* | 194.74819709 | -7.132 | -8.525 | -5.739 |
| **Hedge's correction** | 194.74819709 | -7.039 | -8.414 | -5.664 |
| **Glass's delta** | 242.62033979 | -5.725 | -6.707 | -5.235 |

a. The denominator used in estimating the effect sizes. Cohen's *d* uses the pooled standard deviation. Hedge's correction uses the pooled standard deviation, plus a correction factor. Glass's delta uses the sample standard deviation of the control (i.e., the second group). CI: Confidence interval, SE: Standard error, SD: Standard Deviation

**Figure 2:**
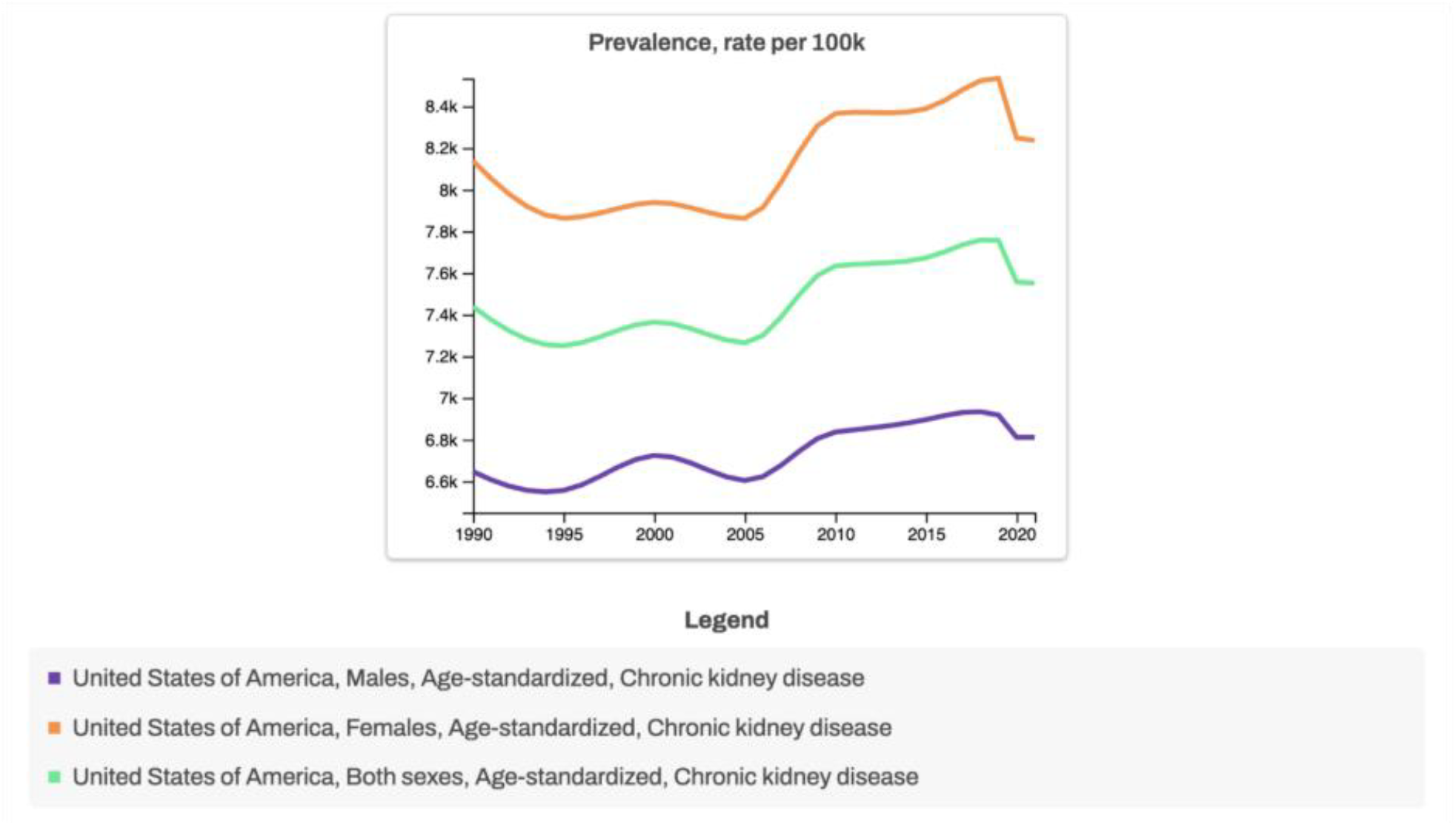

Over the 27-year period, the prevalence of CKD for both sexes slightly increased. In 1990, the prevalence rate for males was 6646.65 per 100,000, increasing to 6917.83 per 100,000 by 2019, a 4.1% increase. For females, the prevalence rate in 1990 was 8138.19 per 100,000, which slightly increase to 8534.31, representing 3.8% increase. Although both sexes experienced increases in prevalence, females consistently exhibited higher amounts of prevalence rates than males throughout the study period.

Temporal trend analysis utilizing linear regression models have also demonstrated a significant increase in the prevalence rates for both males and females. The rate of increase was significantly faster for females (β = 22.463, P < 0.001) compared to males (β = 13.359, P < 0.001) [Table 2]. In short, this finding represents even though females often experienced higher prevalences rates due to CKD, the gap between males and females increased over time.

### Mortality

The mortality of CKD showed significant sex differences [Figure 3]. Males had a higher mean mortality rate (16.94 per 100,000, standard deviation [SD] = 5.15) compared to females (12.35 per 100,000, standard deviation [SD] = 3.81). The independent samples t-test indicated a significant difference in the mortality rates between the sexes (t[58] = 3.918, P < 0.001). Effect size of the metrics also details the magnitude of this difference, with Cohen’s d calculated at 4.53, Hedge’s g at 4.53, and Glass’s Δ at 3.81 [Table 3]. These large effect sizes illustrate that the disparity in CKD mortality rate between males and females was substantial.

**Table 3.** Descriptive Group Statistics of Mortality of Chronic Kidney Disease by Sex

| Sex | n | Mean | SD | SE |
| --- | --- | --- | --- | --- |
| Male | 30 | 16.939801 | 5.154893 | 0.941150 |
| Female | 30 | 12.353443 | 3.811769 | 0.695931 |

**Levene's Test** ***t*-Test of Incidence of Chronic Kidney Disease by Sex**
| Condition | *F* | Significant | *t* | *df* | One-sided (P) | Two-sided (P) | Mean difference | SE difference | Lower CI (95%) | Upper CI (95%) |
| --- | --- | --- | --- | --- | --- | --- | --- | --- | --- | --- |
| Equal variances assumed | 3.014 | =0.088 | 3.918 | 58.000 | <0.001 | <0.001 | 4.58635729 | 1.17050562 | 2.243336 | 6.929379 |
| Equal variances not assumed |  |  | 3.918 | 53.414 | <0.001 | <0.001 | 4.58635729 | 1.17050562 | 2.239045 | 6.933669 |

Independent Sample Effect Sizes for Mortality of Chronic Kidney Disease by Sex
| Test | Standardizer<sup>a</sup> | Point estimate | Lower CI (95%) | Upper CI (95%) |
| --- | --- | --- | --- | --- |
| Cohen's d | 4.53334879 | 1.012 | 0.473 | 1.550 |
| Hedge's correction | 4.53334879 | 0.999 | 0.467 | 1.530 |
| Glass's delta | 3.81176903 | 1.203 | 0.628 | 1.953 |

*a. The denominator used in estimating the effect sizes. Cohen's d uses the pooled standard deviation. Hedge's correction uses the pooled standard deviation, plus a correction factor. Glass's delta uses the sample standard deviation of the control (i.e., the second group). CI: Confidence interval, SE: Standard error, SD: Standard Deviation*

**Figure 3:**
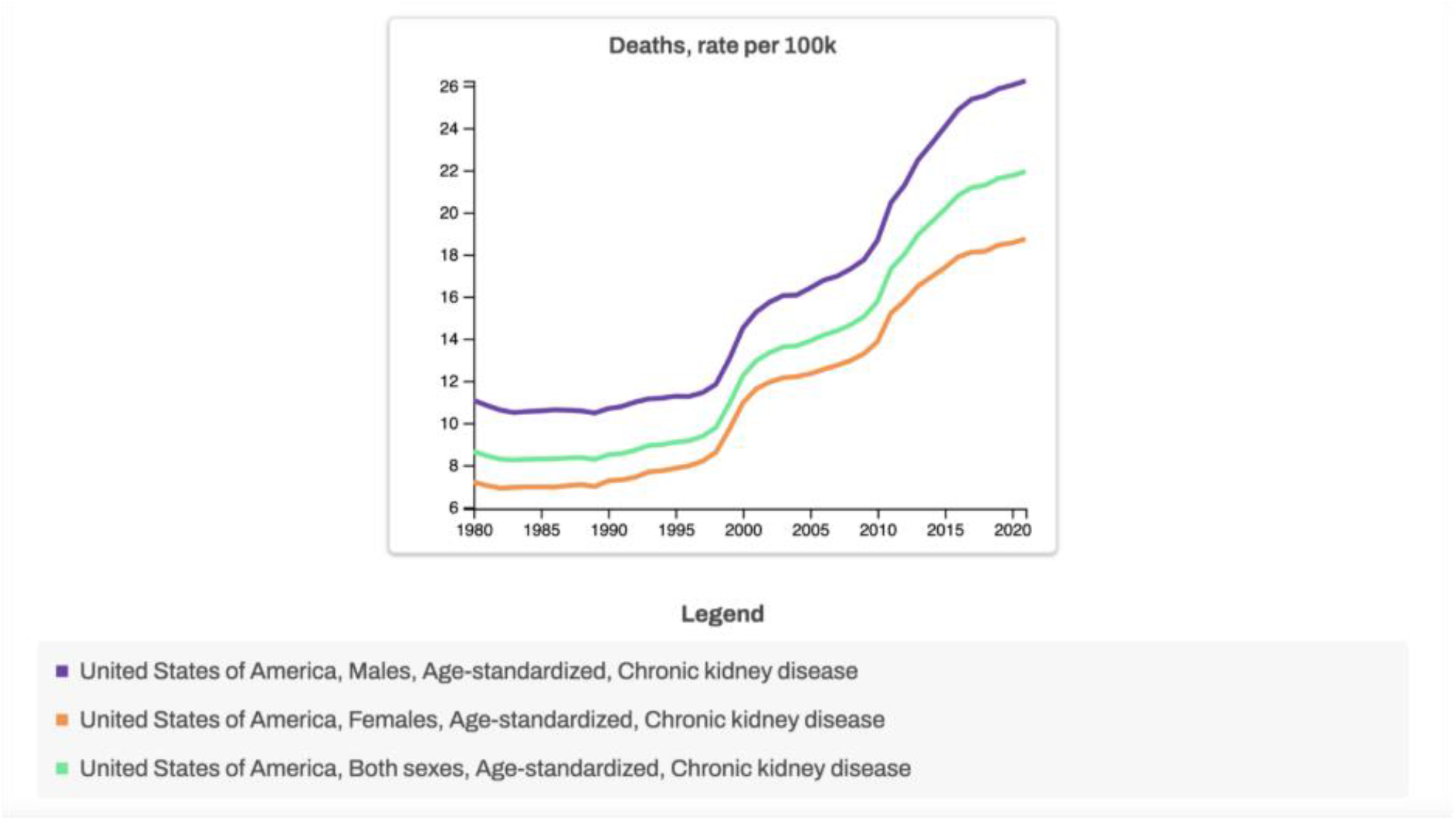

Over the 29-year period, the mortality of CKD for both sexes slightly increased. In 1990, the mortality rate for males was 10.68 per 100,000, increasing to 25.86 per 100,000 by 2019, a 142.13% increase. For females, the mortality rate in 1990 was 7.25 per 100,000, which slightly increase to 18.45, representing 154.48% increase. Although both sexes experienced increases in mortality, males consistently exhibited higher amounts of mortality rates than females throughout the study period.

Temporal trend analysis utilizing linear regression models have also demonstrated a significant increase in the prevalence rates for both males and females. The rate of increase was slightly faster for males (β = 0.573, P < 0.001) compared to females (β = 0.427, P < 0.001) [Table 3]. In short, this finding represents even though males often experienced higher mortality rates due to CKD, the gap between males and females increased over time.

### Temporal Trends Analysis

From the study, the temporal trends in the prevalence and mortality for both sexes suggest an overall increase in CKD outcomes over the 29-year period, experiencing steady increase from 1990 to 2019. Females showed higher rates in both incidence and prevalence, whereas males demonstrated higher rates in mortality, indicating persistent sex differences in PRI outcomes.

Even though both sexes have experienced rises in CKD burden, the sex gaps have widened over time, where females have shown faster increase in prevalence, yet males demonstrated sharper increases in mortality, understanding both persistent and growing disparities in the United States.

## Discussion

This study uncovers the significant sex differences in the incidence, prevalence, and mortality rates because of CKD in the United States from 1990 to 2019. The results demonstrate how females consistently showed higher rates of incidence and prevalence, whereas males showed higher outcomes of mortality of CKD.

Biological mechanisms (i.e., reno-protective effects of estrogen, potential pro-fibrotic actions of androgens, and sex-biased gene expression) may contribute to slower decline in renal function among premenopausal women yet a relatively faster progression in men [9-12]. Moreover, the renal structural and physiological differences (e.g., differences in glomerular size, single-nephron filtration, blood pressure regulation, and immune responses) can further support the different sex diseases paths even when exposed to the same risks such as diabetes or hypertension [10–12].

Healthcare access further shapes these sex differences. Although women show higher rates of incidence and prevalence, they are less likely than men to have their CKD formally recognized or managed after abnormal kidney function is detected. Studies report that women receive fewer follow-up tests, fewer urine albumin checks, and are prescribed renin–angiotensin inhibitors and statins at lower rates [13–15]. When CKD progresses, women are also referred for dialysis and transplantation less often and later than men, despite making up most living kidney donors [14–16]. As a result, women are more visible in early-stage CKD statistics, but men, who experience faster progression and more access to advanced therapies, can explain why they are more likely to appear in dialysis populations and mortality data.

The temporal trends found in this study also provide some important insights. We observed no statistically significant changes in incidence, but prevalence increased steadily in both sexes, while faster in women. This could likely reflect greater survival with CKD, an aging population, and improved detection, specifically within women [1, 9–11]. However, mortality increases significantly within both sexes, slightly faster in men. This aligns with global evidence indicate that men consistently faced higher CKD related mortality, despite improvements in their disease management [1,9,11,]. Together, these patterns indicate that improved detection without equal access to the effective treatment could expand population living with CKD, leaving sex gaps in outcomes unresolved.

These results highlight several priorities for intervention. First, detection in women should be improved by providing routine albuminuria screening, consistent monitoring, and equitable use of proven therapies [13–15]. Second, prevention of progression in men should be emphasized through aggressive control of diabetes, hypertension, and lifestyle risk factors, combined with earlier referral to nephrology care [9–11,15]. Finally, research and quality improvement should consistently stratify outcomes by sex and gender, include menopausal status where possible, and test interventions designed to address these sex-specific drivers of disease [9–11,15,16].

This study has several limitations. Even though the Global Burden of Disease dataset provides comprehensive nation estimates, errors because of underreporting or misclassification could have occurred. Individual-level factors, such as socioeconomic status, race and ethnicity, and mediation use were not available, which may influence the observed trends. Future research should further incorporate biomarkers such as cystatin C and more detailed demographic data to clarify the extent of sex differences in CKD [1, 9–11].

Our study reveals the significant sex differences in CKD in the United States, with females in the United States consistently showing higher incidence and prevalence of CKD, while males faced higher mortality rates. Over time, the prevalence gaps widen in women, while mortality gaps widen in men, highlighting to growing disparities. These findings likely reflect a combination of biological sex differences and gender related differences in healthcare access. Future work should investigate both biological and gendered drivers of these trends to design interventions that reduce sex-based inequities in CKD outcomes.

## Data Availability

All data produced are available online at Global Burden of Disease (GBD)

https://www.healthdata.org/research-analysis/gbd

## References

1. Centers for Disease Control and Prevention. Chronic Kidney Disease in the United States, 2023. Atlanta (GA): US Department of Health and Human Services; 2023 [cited 2025 Sep 28]. Available from: https://www.cdc.gov/kidneydisease/publications-resources/ckd-national-facts.html

2. National Center for Health Statistics. Final Data on Leading Causes of Death, 2022. Hyattsville (MD): Centers for Disease Control and Prevention; 2025.

3. National Institute of Diabetes and Digestive and Kidney Diseases. Kidney Disease Statistics for the United States. Bethesda (MD): National Institutes of Health; 2024.

4. National Kidney Foundation. CKD Causes and Risk Factors. New York (NY): National Kidney Foundation; 2024.

5. National Kidney Foundation. Kidney Disease Fact Sheet. New York (NY): National Kidney Foundation; 2024.

6. United States Renal Data System. 2023 Annual Data Report: Epidemiology of Kidney Disease in the United States. Bethesda (MD): National Institute of Diabetes and Digestive and Kidney Diseases, National Institutes of Health; 2023.

7. Ricardo AC, Yang W, Sha D, et al. Sex-related disparities in CKD progression. J Am Soc Nephrol. 2019;30(1):137–46. doi:10.1681/ASN.2018030296

8. Ahmed SB, Dumanski SM. Do sex and gender matter in kidney disease? Am J Kidney Dis. 2021;78(2):176–9. doi:10.1053/j.ajkd.2021.03.335

9. Carrero JJ, Hecking M, Chesnaye NC, Jager KJ. Sex and gender disparities in the epidemiology and outcomes of chronic kidney disease. Nat Rev Nephrol. 2018;14(3):151–64. doi:10.1038/nrneph.2017.181

10. Franco-Acevedo A, Rojas-Villarraga A, Espinosa G. Sex differences in renal function: hormones and prolactin. Endocrines. 2021;2(4):460–74. doi:10.3390/endocrines2040037

11. Hockham C, Newton R, Woodward M. Sex differences in chronic kidney disease mortality: insights from the Global Burden of Disease Study 1990–2019. Kidney Med. 2022;4(5):100412. doi:10.1016/j.xkme.2022.100412

12. Neugarten J, Golestaneh L. Influence of sex on the progression of chronic kidney disease. Mayo Clin Proc. 2019;94(7):1339–56. doi:10.1016/j.mayocp.2018.10.021

13. Swartling O, Evans M, Szummer K, et al. Sex differences in the recognition, monitoring, and management of CKD in health care. J Am Soc Nephrol. 2022;33(9):1753–64. doi:10.1681/ASN.2021101377

14. Smothers L, Pastan S, Mohottige D, et al. Gender disparities in early referral for kidney transplantation: results from a Southeast U.S. cohort. Kidney Int Rep. 2022;7(2):246–54. doi:10.1016/j.ekir.2021.10.015

15. Tong A, Sainsbury P, Chadban S, et al. Gender disparities in access to kidney transplantation: perspectives of nephrologists. Kidney Int Rep. 2022;7(7):1674–83. doi:10.1016/j.ekir.2022.04.096

16. Mayne KJ, Hockham C, Woodward M, et al. Sex and gender differences in chronic kidney disease management: a narrative review. J Hum Hypertens. 2023;37:409–18. doi:10.1038/s41371-022-00797-2

